# Kir2.1 mutations differentially increase the risk of flecainide proarrhythmia in Andersen Tawil Syndrome

**DOI:** 10.1101/2024.12.10.24318629

**Authors:** Francisco M. Cruz, Ana I. Moreno-Manuel, Sánchez Pérez Patricia, Juan Manuel Ruiz-Robles, Paula García Socuellamos, Lilian K. Gutiérrez, María Linarejos Vera-Pedrosa, Amaia Talavera Gutierrez, Gema Mondéjar Parreño, Álvaro Macías, Isabel Martínez-Carrascoso, Francisco J Bermúdez-Jiménez, Salvador Arias Santiago, Fernando Martínez de Benito, Aitana Braza-Boils, Carmen Valenzuela, CA Morillo, Esther Zorio, Juan Jiménez-Jaimez, José Jalife

## Abstract

**Background:** Flecainide and other class-Ic antiarrhythmic drugs (AADs) are widely used in Andersen-Tawil syndrome type 1 (ATS1) patients. However, class-Ic drugs might be proarrhythmic in some cases. We investigated the molecular mechanisms of class-I AADs proarrhythmia and whether they might increase the risk of death in ATS1 patients with structurally normal hearts.

**Methods and Results:** Of 53 ATS1 patients reviewed from the literature, 54% responded partially to flecainide, with ventricular arrhythmia (VA) reduction in only 23%. Of the latter patients, VA persisted in 20-50%. Flecainide was ineffective in 23%, and surprisingly, 13.5% suffered a non-fatal cardiac arrest. In five cardiac-specific ATS1 mouse models (Kir2.1^Δ314–315^, Kir2.1^C122Y^, Kir2.1^G215D^ and Kir2.1^R67W^ and Kir2.1^S136F^), flecainide or propafenone (40 mg/Kg i.p.) differentially prolonged the P wave, and the PR, QRS and QTc intervals compared to Kir2.1^WT^; Kir2.1^S136F^ had milder effects. Flecainide increased VA inducibility in all mutant mice except Kir2.1^S136F^, which exhibited significant VA reduction. At baseline, Kir2.1^G215D^ cardiomyocytes had the lowest inward rectifier K+ channel (I_K1_) reduction, followed by Kir2.1^C122Y^, Kir2.1^R67W^ and Kir2.1^S136F^. Kir2.1^C122Y^ cardiomyocytes had a significant decrease in sodium inward current (I_Na_). Flecainide (10 µM) slightly increased I_K1_ density in Kir2.1^WT^ and Kir2.1^S136F^, while it decreased both I_K1_ and I_Na_ in Kir2.1^C122Y^ and Kir2.1^R67W^, despite normal trafficking of mutant channels. Optical mapping in ATS1 patient-specific iPSC-CM monolayers expressing Kir2.1^C122Y^, Kir2.1^G215D^ and Kir2.1^R67W^ showed an increase in rotor incidence at baseline and under flecainide, confirming the druǵs proarrhythmic effect. Lastly, in-silico molecular docking predicts that the Kir2.1-Cys_311_ pharmacophore-binding site is altered in Kir2.1^C122Y^ heterotetramers, reducing flecainide accessibility and leading to channel closure and arrhythmias.

**Conclusions:** Class-Ic AADs are only partially effective and might be proarrhythmic in some ATS1 patients. Kir2.1 mutations impacting the resting membrane potential and cellular excitability create a substrate for life-threatening arrhythmias, raising significant concern about using these drugs in some ATS1 patients.

**CLINICAL PERSPECTIVE NOVELTY AND SIGNIFICANCE:** *What is known?:*

- Andersen-Tawil syndrome type 1 (ATS1) is a rare autosomal dominant disease caused by loss-of-function mutations in the *KCNJ2* gene, which encodes the Kir2.1 channel responsible for the repolarizing, strong inwardly rectifying current I_K1_.
- ATS1 treatment is empirical and subject to clinical judgment. It includes the use of class-Ic antiarrhythmic drugs (AADs), mainly flecainide, alone or in combination with β-adrenergic blocking drugs. However, pharmacological treatment is partial and might fail, leading to life-threatening ventricular arrhythmias (VA) and sudden cardiac death (SCD) in some ATS1 patients.
- Some ATS1 mutations are known to disrupt the Kir2.1-Nav1.5 channelosome in mice and human iPSC-CMs, with consequent reductions in cardiac excitability and conduction velocity (CV), leading to VA, which may be exacerbated by flecainide.

*What new information does this article contribute?:*

- In our analysis of 53 ATS1 patients, flecainide showed partial effectiveness. While a few patients experienced complete disappearance of VA, others had persistent arrhythmias and even suffered non-fatal cardiac arrest while on medication.
- In murine models expressing five relevant ATS1 mutations, flecainide or propafenone produced differential alteration in the P wave, PR, QRS and QTc intervals, and increased VA inducibility compared with Kir2.1^WT^ mice. Additionally, flecainide differentially affected I_K1_ and the Na^+^ inward current (I_Na_) current densities despite normal trafficking of mutant channels.
- In patient-specific induced pluripotent stem cell derived cardiomyocyte (iPSC-CM) monolayers flecainide reduced CV and increased rotor incidence, confirming the drugś proarrhythmic effect.
- *In-silico* molecular docking studies predicted that the Cys_311_ pharmacophore binding site and flecainide accessibility are altered in mutated Kir2.1 channels, leading to premature channel closure and arrhythmias.
- We conclude that class-Ic AADs are only partially effective and might be proarrhythmic in some ATS1 patients.
- These findings raise concern about the use of class-Ic AADs in ATS1 patients and highlight the need for further studies to guide personalized therapy.

## Introduction

Andersen-Tawil syndrome type 1 (ATS1) is a rare autosomal dominant disease caused by loss-of-function mutations in the *KCNJ2* gene, which codes the strong inward rectifier potassium channel Kir2.1 responsible for the repolarizing current IK1.^1^ IK1 plays a vital role maintaining the resting membrane potential (RMP) and the final phase of action potential (AP) repolarization. Thus, loss-of-function mutations in *KCNJ2* substantially decrease I_K1_, with consequent prolongation of the AP duration (APD) and corrected QT (QTc) interval.^2^ These events predispose ATS1 patients to a triad of symptoms including periodic paralysis, dysmorphias, and cardiac electrical alterations that may lead to high-burden polymorphic ventricular extrasystoles, biventricular tachycardia (BiVT), and sudden cardiac death (SCD)^3^ by mutation-dependent mechanisms that are far from being fully understood. Accordingly, it is unknown why life-threatening arrhythmias occur in these patients, why some mutations have a more severe phenotype than others, even within the same family, and how to treat each individual patient. Currently ATS1 treatment is empirical and subject to clinical judgment.^4^ Medical devices like implantable cardiac defibrillators (ICDs) are frequently indispensable in the management of severe and symptomatic cardiac arrhythmias. However, ICDs may induce inappropriate shocks and patients are not exempt from SCD risk.^4–6^ Pharmacological treatment includes class-Ic antiarrhythmic drugs (AADs), mainly flecainide or propafenone alone or in combination with β-adrenergic blocking drugs.^7,8^ However, class-Ic drugs may fail or are proarrhythmic in ATS1 patients^4^ due to poorly understood mechanisms.

More than 90 ATS1-associated loss-of-function mutations have been identified and distributed throughout the Kir2.1 protein structure. Mutations may affect channel trafficking, channel gating, or ion selectivity properties through molecular mechanisms that are incompletely understood.^9^ Additionally, Kir2.1 forms channelosomes with Na_V_1.5 proteins, responsible for the sodium inward current (I_Na_).^10–12^ This specialized molecular complex plays a crucial role in maintaining cardiac excitability and rhythm by stabilizing the RMP and initiating the AP, ensuring coordinated cardiac activity. We have previously shown that, in addition to sequestering the Kir2.1 protein in the Golgi apparatus,^13^ the Kir2.1^Δ314–315^ mutation also prevents Na 1.5 membrane trafficking, thus altering the function of both proteins *in-vitro* and *in-vivo*.^20^ More recently, we demonstrated that, while maintaining trafficking ability, the extracellular Kir2.1^C122Y^ mutation that disrupts interactions of the channel with phosphatidyl inositol 4,5-bisphosphate (PIP_2_) also produces temporal instability of both Kir2.1 and Na_V_1.5 proteins and reduces excitability *in-vivo*.^14^ Thus, the above results underscore the complexity of the mechanisms governing Kir2.1-Na_V_1.5 channelosome function, and it is clear that each ATS1 mutation exhibits distinct molecular mechanisms despite targeting the same gene. To date, only a handful of studies have addressed the potential alterations in the surrounding molecular context that governs cardiac activity, including Na_V_1.5 channels. There is, therefore, a significant discrepancy between what is known and what should be known regarding molecular mechanisms for a more effective treatment of each specific mutation leading to ATS1^15,16^. The lack of mechanistic knowledge significantly hinders our understanding of the cardiac arrhythmia phenotype and prevents progress in disease treatment and SCD prevention. No studies have been conducted to date to determine the impact of flecainide or any other class-Ic AADs on the electrical activity of the Kir2.1-Na_V_1.5 channelosome, which could vary significantly depending on the specific ATS1 mutation.

In this article, we review available clinical data and demonstrate that the cardiac electrical response of ATS1 patients to class-Ic AADs is highly variable, and at times contrary to anticipated. While some patients benefit from the use of these drugs, others do not, and yet others may get worse. We hypothesize that the ultimate effect of class-Ic AADs in a given ATS1 patient will depend on the precise electrophysiological consequences of the Kir2.1 loss-of-function mutation. Specifically, we surmise that mutations impacting both RMP (I_K1_ reduction) and cellular excitability by altering Na_V_1.5 function (I_Na_ reduction) and channel availability (through RMP-mediated depolarization), may potentially create a substrate for life-threatening arrhythmias, particularly promoted by class-Ic AADs treatment. Our approach is multidisciplinary, and our results emphasize the need to elucidate the pathophysiology of arrhythmias in individual patients with ATS1, and identify an effective, more personalized therapy that reduces proarrhythmia risk.

## Materials & Methods

*The authors declare that all supporting data are available within the article. Please see the Major Resources Table and Supplemental Methods for more details*.

### Ethics Statement

All animal experimental procedures conformed to EU Directive 2010/63EU and Recommendation 2007/526/EC. We obtained skin biopsies from patients carrying the Kir2.1^C122Y^, Kir2.1^G215D^ and Kir2.1^R67W^ mutation after written informed consent, previously approved by local institutional review (2020-411-1, 2020-582-1), according to the Ethics Committee for Research of CNIC and the Carlos III Institute (CEI PI58_2019-v3), Madrid, Spain. The local ethics committees and the Animal Protection Area of the Comunidad Autónoma de Madrid (PROEX 111.4/20 and 226.5/23) approved the animal protocols.

### Mice

We obtained 4-5-weeks-old C57BL/6J male mice from the Charles River Laboratories. Animals were reared and housed according to CNIC animal facility guidelines and regulations.

### AAV vector production, purification, and mouse model generation

AAV vectors were generated using the cardiomyocyte-specific troponinT proximal promoter (cTnT) and encoding wildtype Kir2.1 (Kir2.1^WT^) or the ATS1 mutant channels (Kir2.1^Δ314–315^, Kir2.1^C122Y^, Kir2.1^G215D^ and Kir2.1^R67W^ and Kir2.1^S136F^), followed by tdTomato report. Vectors were packaged into AAV serotype 9 (AVV9) and produced by the triple transfection method, using HEK293T cells as described previously.^17,18^ Mice were anesthetized with ketamine (60 mg/kg) and xylazine (20 mg/kg) by intraperitoneal (i.p.) route. Thereafter, mice were inoculated with 3.5^x^10^10^ viral particles through the femoral vein at a final volume of 50μL. Only well-inoculated animals were included in the studies. All experiments were performed 8-to-10 weeks after infection. *Ex-vivo* fluorescent signal confirming cardiac expression and distribution of protein expression was assessed as described.^19^

### Drugs

Flecainide-acetate (10mg/ml injection solution; Mylan pharmaceuticals) and propafenone-hydrochloride (Sigma) were dissolved and administered i.p. in mice at a single dose of 40 mg/kg.^20^ Fresh drug solutions were prepared for each experiment and further dilutions were carried out in external solution to obtain the desired final concentration for patch clamping and optical mapping experiments. Control solutions always contained the same solvent concentrations as the test solution. ECGs were recorded for 1 min at baseline and an additional 5 min after intraperitoneal injection of flecainide or propafenone. hiPSCs-CM monolayers were incubated in the presence of 1 μM flecainide and isolated mouse cardiomyocytes expressing Kir2.1^WT^, Kir2.1^C122Y^, Kir2.1^R67W^, Kir2.1^G215D,^ Kir2.1^R67W^ and Kir2.1^S136F^ channels were recorded at baseline and after 10 min of incubation with a final concentration of 10 μM of flecainide (IC_50_ for Na_V_1.5 channels).^21^

### Surface ECG recording

Mice were anesthetized using isoflurane inhalation (0.8-1.0% volume in oxygen) and maintained at 37°C on a heating plate. Four-lead surface ECGs were recorded for 6 min (1 min baseline + 5 min after i.p. drug) using subcutaneous limb electrodes connected to an MP36R amplifier unit (BIOPAC Systems). Data acquisition and analyses were performed using AcqKnowledge software.

### *In-vivo* intracardiac recording, stimulation, and drug administration

An octopolar catheter (Science) was inserted through the jugular vein and advanced into the right ventricle as previously described.^22^ VA inducibility was assessed by applying consecutive trains at 10Hz and 25Hz, respectively.

### Cardiomyocyte isolation

The procedure was performed as previously described.^23^ (See *Supplemental Methods)*.

### Patch-clamping in isolated cardiomyocytes

The whole-cell patch-clamp technique as well as data analysis procedures and internal and external solutions (***Supplementary Table 1***) were similar to those previously described.^9–13^ Details are presented in the *Supplemental Methods*.

### Generation, culture, and differentiation of iPSC-CMs

We reprogrammed primary fibroblasts derived from skin biopsies from ATS1 patients (C122Y, R67W and G215D) to iPSCs using the Sendai virus for transfection of Yamanaka’s factors: OCT4, KLF4, c-Myc, and SOX2, as previously described.^24^ HiPSCs were differentiated into hiPSC-CMs using small molecules-mediated canonical Wnt pathway modulation as previously described.^25,26^ HiPSC-CMs were purified from non-myocytes following the MACS negative selection protocol (Miltenyi Biotec Kit) and seeded for 7 days on a PDMS substrate to improve cardiomyocyte maturation. See *Supplemental Methods* for details.

### Optical Mapping in iPSC-CMs monolayers

Phase, conduction velocity and activation maps were generated at baseline and after pacing (from 1000-to-400 ms). Flecainide (5 uM) was incubated for 1 min before pacing, and experiments were carried out as previously described.^27,28^.

### Statistical analyses

To determine the statistical power and minimum sample size in our experiments, we did a power calculation using R base “power.t.test” or “power.anova.test” function depending on data analysis. We used a significance level (alfa=0.05), power (1-beta=80%), the estimated difference between control and experimental data for t-test, and estimated variances for ANOVA. We used GraphPad Prism software versions 7.0 and 8.0. For non-Gaussian distributions, we applied the non-parametric Mann-Whitney test. We used one- or two-way ANOVA corrected by Šídák’s multiple comparisons test. We used Grubb’s test for analysis of outliers. Data are expressed as mean ± SEM, and differences are considered significant at p<0.05 (*p<0.05; **p<0.01; ***p<0.001; ****p<0.0001). Note that “N” refers to the number of mice or hiPSC-CMs differentiations used and “n” to the number of cells analyzed per mouse or monolayer.

## Results

### Ventricular arrhythmias persist in ATS1 patients despite flecainide therapy

We systematically scanned the existing ATS1 literature assessing the effects of an empirical, “one-size-fits-all” antiarrhythmic treatment, specially the use of class-Ic AADs flecainide or propafenone, either alone or in combination with β-blockers. Our analysis included both genders, all age ranges and diverse origins, while specifically excluding those with structural or ischemic heart disease. The search yielded a limited number of publications (23), most of which were isolated case reports (∼87%) or small series (less than 12 patients; ∼13%), without any follow-up report. We evaluated the cardiac phenotype, therapy and clinical response in a total of 53 documented patients each carrying one of more than 20 different Kir2.1 loss-of-function mutations (**Table 1**). We incorporate a recently published study^14^ including a Kir2.1^C122Y^ proband, who presented several syncopal polymorphic VT and suffered three proper ICD shocks between ages 25-35 under sodium channel blocker administration. A Holter trace (**Figure 1A**) shows an electrogram with long-short coupling initiating polymorphic VT (PVT) suggesting either ineffective therapy or a druǵs proarrhythmic effect.

**Figure 1.**
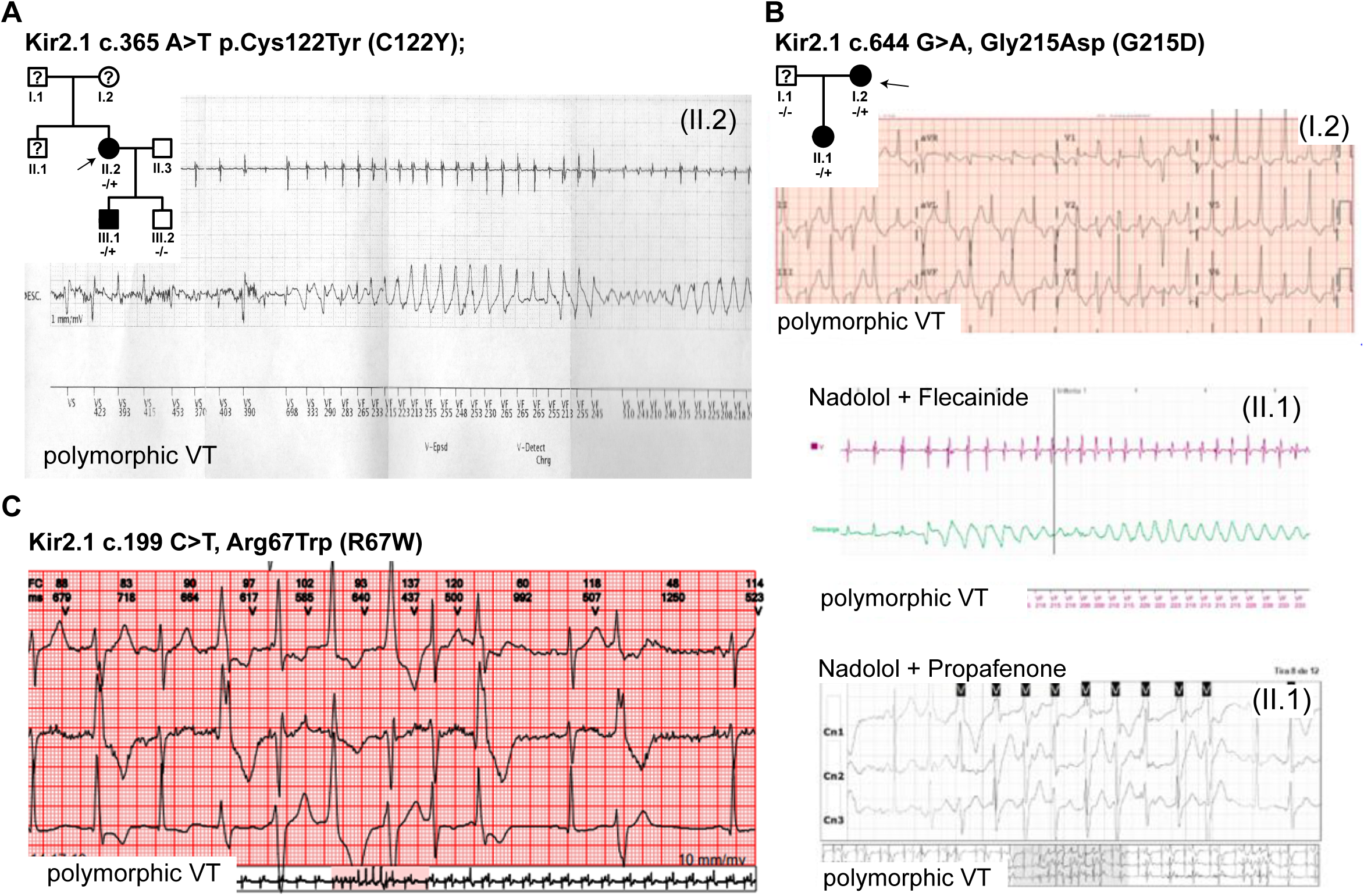
Arrhythmogenic responses to Ic class AADs in ATS1 patients. **A**: Recorded polymorphic ventricular tachycardia (VT) from an automated external defibrillator in Kir2.1^C122Y^ proband (II.2). Pedigree of the family is shown in the upper left square. **B**: Recorded polymorphic ventricular tachycardia (*upper panel*) and a high burden of non-sustained ventricular tachycardias (NSVT) in Kir2.1^G215D^ proband (I.2) under Flecainide + Nadolol (*middle panel*) or Propafenone + Nadolol (*bottom panel*). **C**: ECG from Kir2.1^R67W^ patient shows polymorphic VT. All Probands are indicated with a black arrow.

**Table 1:**
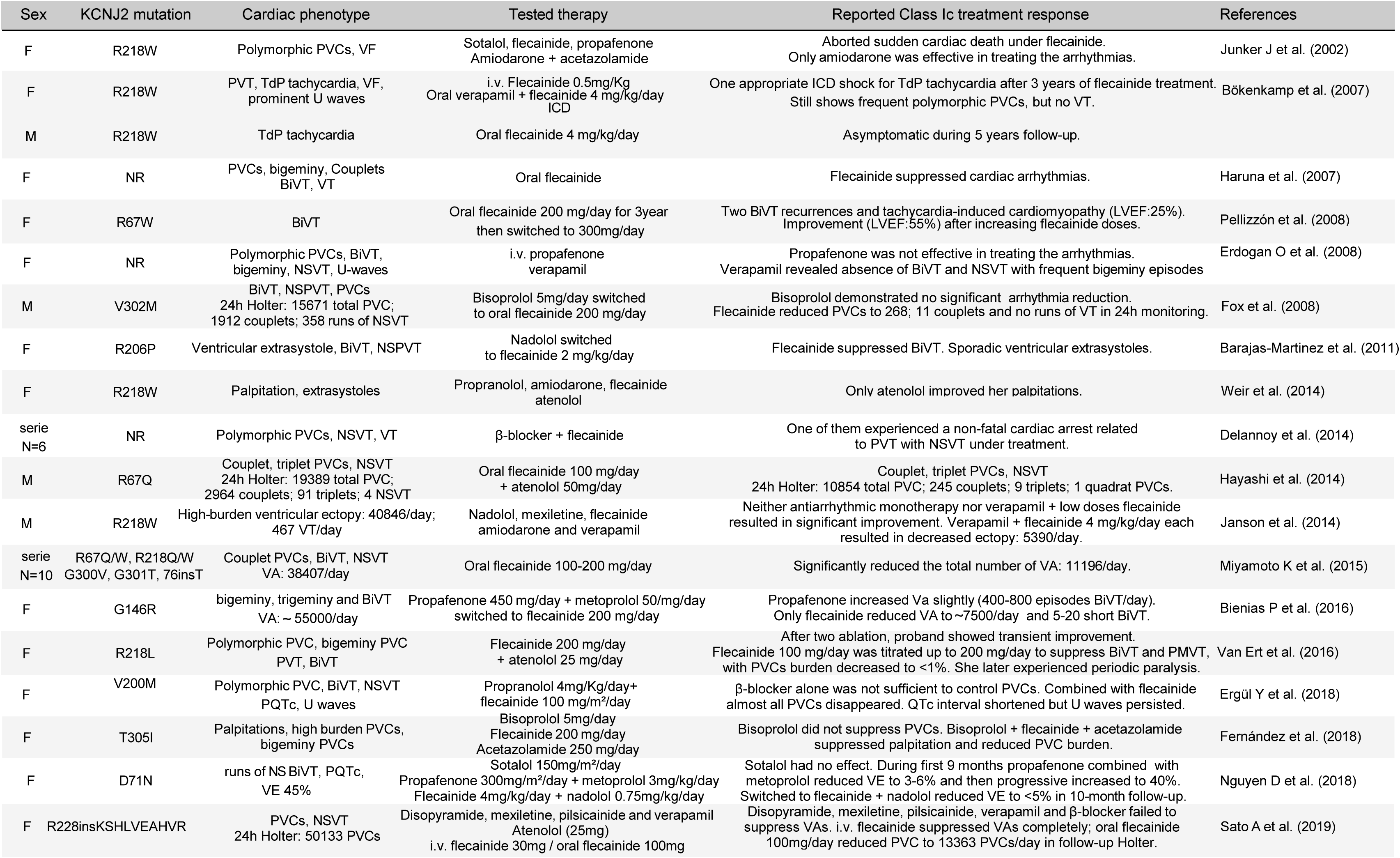
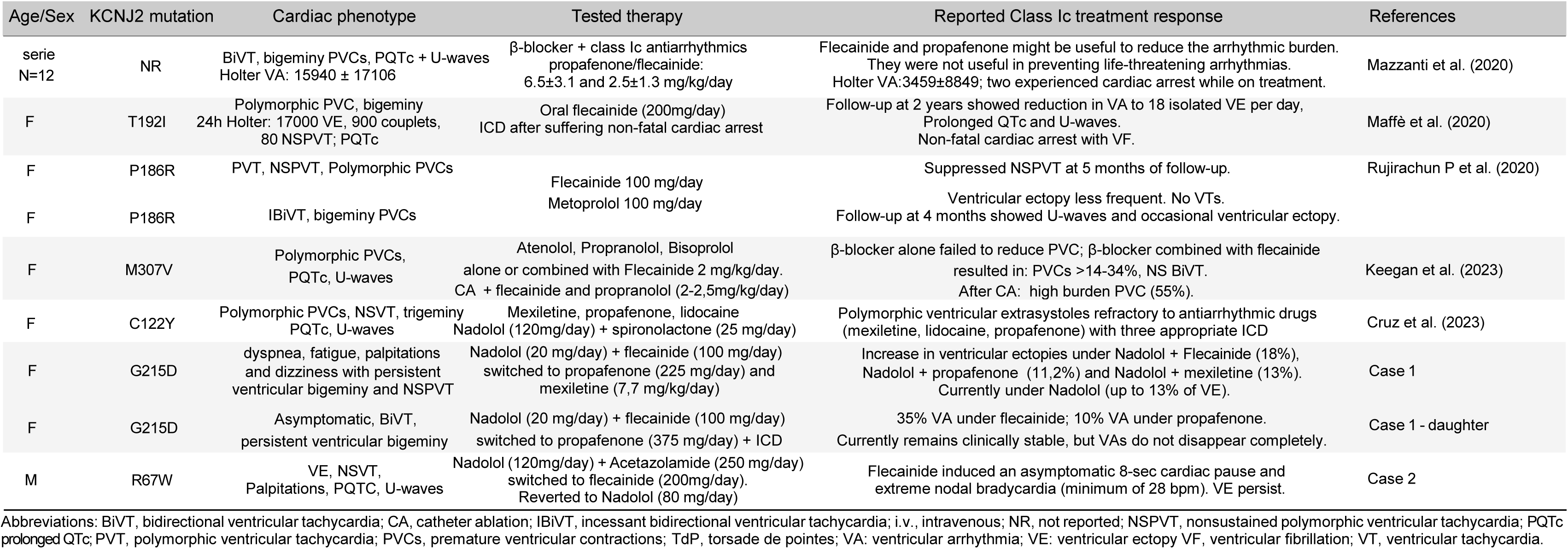
Response to class-Ic AADs in ATS1 patients. Detailed analysis of 53 different probands shows a large variability from patient to patient. Data reveal moderate, incomplete or ineffective responses to class-Ic AADs in suppressing ventricular arrhythmias (VA) in ATS1. The table incorporates demographic information and clinical features of reviewed ATS1 cases, including age, sex, reported mutation, cardiac phenotype, treatment, and response.

We also reproduce previously unreported tracings from two members, mother and daughter, of a Spanish family of Caucasian origin with a final diagnosis of ATS1. Both patients came to the consult of one of the authors (EZ) more than a decade after being studied in other centers and treated ineffectively with multiple drug combinations. Both carried a heterozygous missense *KCNJ2* variant (c.644 G>A, Gly215Asp (G215D)) previously reported to disrupt Kir2.1-PIP_2_ interactions.^29^ The mother, patient I-1, was a female who experienced worsening dyspnea and fatigue in their 60’s (**Case 1 in Table 1**). The ECG showed high burden ventricular extrasystoles (VE) and bidirectional VT (**Figure 1B**, *upper panel*). The patient initially refused β-blocker treatment and was started on flecainide (100 mg per day), but there was no improvement. Flecainide was subsequently combined with small dose nadolol (20mg mg/day). While there was some improvement the patient continued having high burden ventricular extrasystoles (from 3.2 to 18%) and bidirectional VT (**Figure 1B**, *middle panel*). Flecainide was then switched to propafenone (225 mg/kg/day), which led to another hospital admission due to non-syncopal dizziness and continued high VA burden (11.2% with NSVT) (**Figure 1B**, *bottom panel*). The patient experienced intolerance to mexiletine (7.7 mg per day), and after rejecting ICD implantation, she is under nadolol alone, but continues with multiple ventricular extrasystoles (up to 32%) and NSVT (**Figure 1B**) including self-limiting AF (<5 min).

The daughter, patient II-1, was initially referred to the hospital in their 20’s for syncope and multiple VEs (**Case 1-daughter in Table 1**). Nadolol (20mg) combined with flecainide (100 mg/day) partially reduced in VE burden that finally led to a syncope and an ICD was implanted. Treatment was maintained and resulted in a self-limiting VT episode. Medication was changed to nadolol (20mg) and propafenone (375mg per day). She has not reported syncope since then and the NSVT burden has declined from 35% under flecainide to 10% with propafenone. Actually, she remains clinically more stable, but the arrhythmias have not completely disappeared.

We also present the case of an ATS1 male proband in their 40’s carrying the PIP_2_-associated Kir2.1^R67W^ mutation (**Case 2 in Table 1**).^30^ The patient exhibited characteristic dysmorphic features including clinodactyly and micrognathia, and had experienced periodic muscle paralysis since the age of 8, particularly during intense physical activity. Initial treatment consisted of nadolol (120 mg/day) and acetazolamide (250 mg/day). Holter monitoring revealed a QT interval of 480 ms (QTc 434 ms), prominent U waves, high density polymorphic VE and 7 episodes of nonsustained BiVT. The 3-lead ECG time series in **Figure 1C** shows frequent premature ventricular complexes and the Holter fragment at the bottom shows an episode of polymorphic VT. Nadolol was then switched to flecainide (100 mg/12h). However, while NSVT runs were no longer observed in subsequent 24h-Holter monitoring, the patient continued with high density polymorphic VE (>20%) and palpitations. Additionally, flecainide induced an asymptomatic 8-sec cardiac pause and extreme sinus bradycardia, with an average heart rate of 50 bpm and a minimum of 28 bpm. Currently, the patient is on nadolol (80mg/day) and has been scheduled for left cardiac sympathetic denervation.

The Venn diagram presented in **Figure 2** includes the 53 documented patients reported in the 23 publications listed on **Table 1**. Only 23% of patients were successfully treated with complete or greater than 90% VA suppression; 54% presented moderate improvement during class-Ic AADs treatment, maintaining 20-50% of VA. More importantly, in 23% of the cases, flecainide or propafenone were completely ineffective in treating arrhythmias. Surprisingly, a total of 7 patients (13.5%) experienced an additional non-fatal cardiac arrest in the presence of medication. Altogether, the results strongly suggest that the use of flecainide and other class-Ic AADs is debatable in a significant proportion of ATS1 patients due to great variability in outcomes and the potential for life-threatening proarrhythmia. Therefore, the intolerance to class-Ic AADs experienced by many of these patients may depend on the specific ATS1-causative mutation, highlighting the urgent need for molecular studies to investigate the underlying pathophysiological mechanisms.

**Figure 2.**
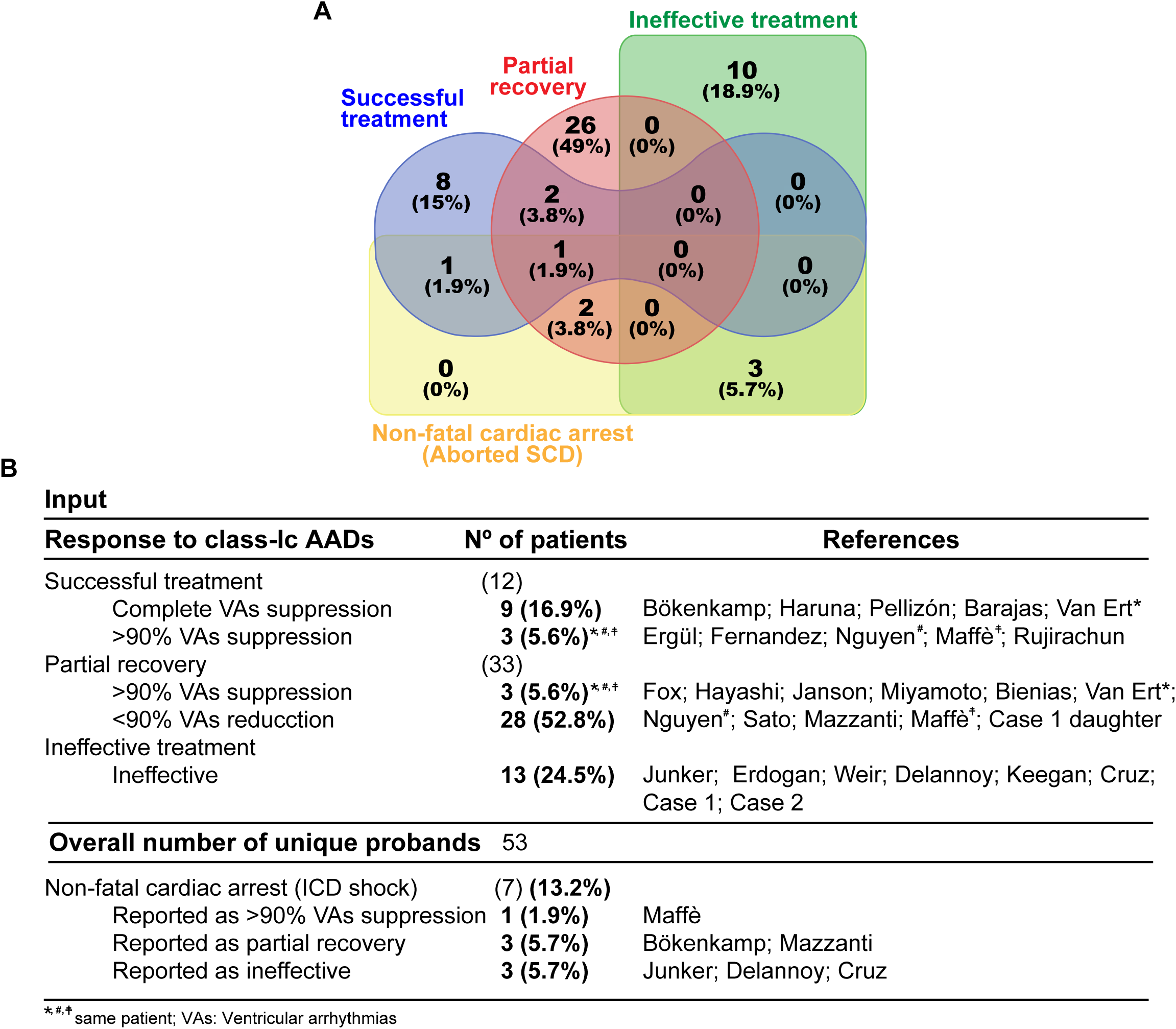
Class-Ic-based antiarrhythmic therapy in AST1. **A-B**: Venn diagram (**A**) and schematic representation (**B**) of class-Ic therapy response in ATS1 patients. Data show 23% of patients who benefited from a full or greater than 90% suppression of VA; 54% reported substantial but incomplete improvement during flecainide treatment, mostly maintaining 20 to 50% of VA. In 23% of patients, flecainide was completely ineffective; 13.5% experienced an additional non-fatal cardiac arrest with an appropriate ICD shock in the presence of flecainide.

### Class-Ic AADs alter cardiac conduction in ATS1 mouse models

The foregoing data demonstrate that the response of ATS1 patients to conventional AADs therapy is highly variable and may depend on the specific mutation and other intermediate factors that affect the patient’s electrical phenotype. To gain insight into what such factors may be, we have generated five relevant cardiac-specific ATS1 mouse models using well-established AAV technology.^19^ In addition to wildtype (Kir2.1^WT^) mice, we generated mice with the cardiac-specific trafficking deficient Kir2.1^Δ314–315^ mutation and the mutation Kir2.1^C122Y^ that disrupts Kir2.1-PIP_2_ interaction, both affecting Kir2.1-Na_V_1.5 channelosomes and excitability.^14,23^ We also included mice with the PIP_2-_related Kir2.1^G215D^ and Kir2.1^R67W^ mutations found in our ATS1 patients. Further, we incorporated the unrelated Kir2.1^S136F^, which has been shown to alter the K^+^ selectivity filter by modifying the highly conserved GYG motif.^9,31,32^ Surface ECG recordings obtained 8 weeks after AAV9-mediated inoculation showed that, compared with Kir2.1^WT^, a single dose of flecainide (40 mg/kg; i.p.)^20^ progressively prolonged the P wave, PR interval, and QRS complex duration in all mutant mice over ∼6 min. However, the effects were different depending on the mutation, with Kir2.1^R67W^ > Kir2.1^G215D^ > Kir2.1^Δ314–315^ > Kir2.1^C122Y^ > Kir2.1^S136F^ (**Figure 3A and Supplemental Figure 1**). These data suggest that the varying effects of each mutation reducing Kir2.1 and Na_V_1.5 membrane function, together with drug-induced I_Na_ blockade synergized to prolong mutation-dependent intra-atrial (P-wave) atrioventricular (PR interval) and intraventricular (QRS) conduction. Additionally, all mutations prolonged the QTc interval to varying degrees, suggesting a differential flecainide-mediated I_K1_ response among mutant channels (Kir2.1^C122Y^ > Kir2.1^G215D^ > Kir2.1^Δ314–315^ > Kir2.1^R67W^ > Kir2.1^S136F^). Propafenone (40 mg/Kg; i.p.) also prolonged all ECG parameters compared with WT but to a lesser extent than flecainide, with the following pattern: Kir2.1^Δ314–315^ > Kir2.1^C122Y^ > Kir2.1^G215D^ > Kir2.1^R67W^ > Kir2.1^S136F^ (**Figure 3B)**.

**Figure 3.**
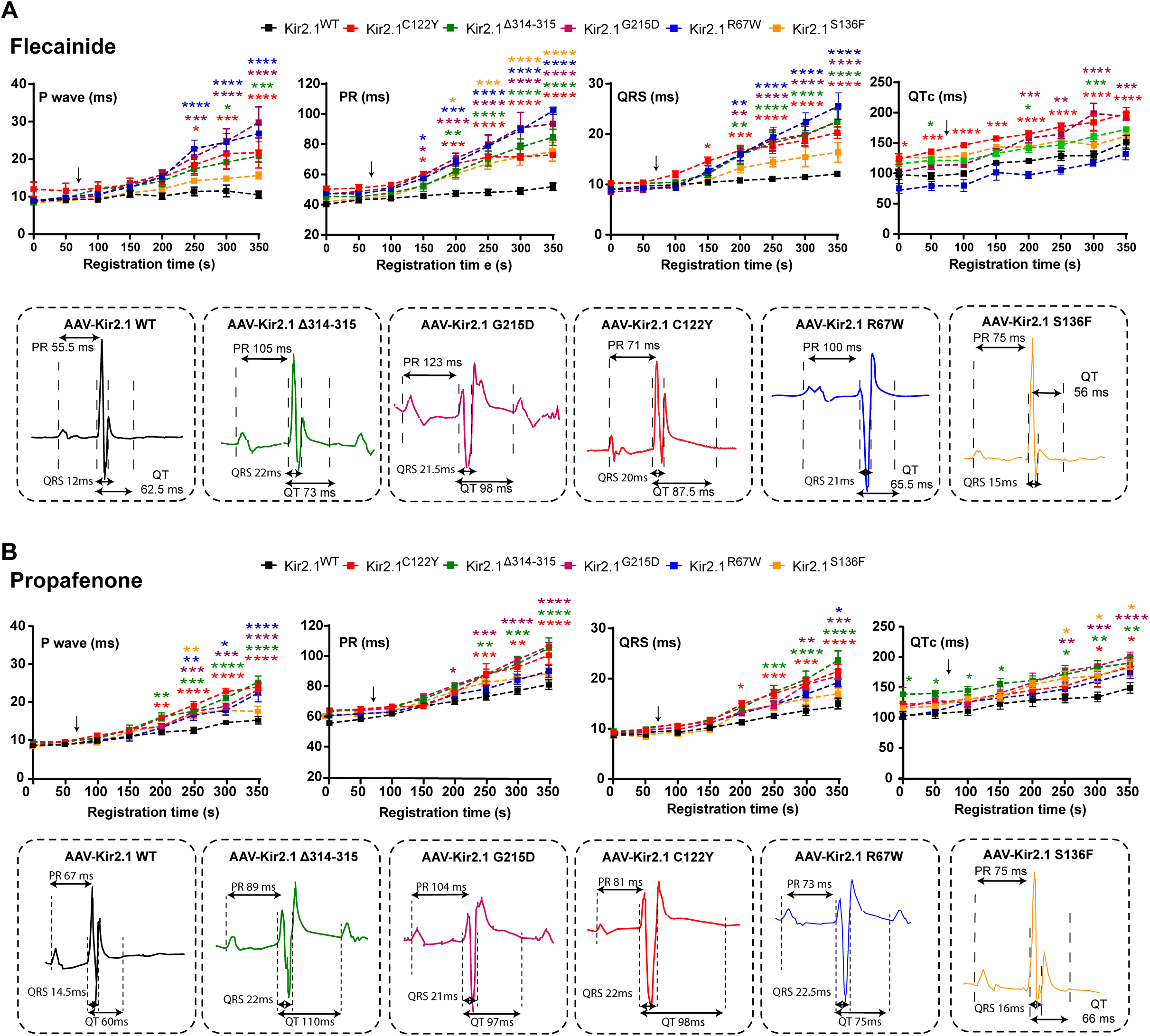
ATS1 mice recapitulate the pathological ECG phenotype. **A-B**: Time-course after a single dose of flecainide **(A)** or propafenone (40 mg/Kg) **(B)** reveals prolonged P wave, PR, QRS and QTc in Kir2.1 mutant animals compared to controls (black). Every value represents the averaged P waves, PR, QRS and QTc intervals from ten consecutive beatings. Arrows indicate the time of flecainide/propafenone administration. Representative beats of Kir2.1^WT^ (black), Kir2.1^Δ314–315^ (green), Kir2.1^C122Y^ (red), Kir2.1^G215D^ (purple), Kir2.1^R67W^ (blue) and Kir2.1^S136F^ (orange) mice after 5 min of flecainide administration are also indicated. Statistical analysis by two-tailed ANOVA. * = p<0.05; ** = p<0.01; **** = p<0.0001.

Analysis of the ECGs revealed that in addition to altering the duration of intervals, many of the Kir2.1^Δ314–315^, Kir2.1^C122Y^, Kir2.1^G215D^ and Kir2.1^R67W^ mice developed spontaneous VA, some of them complex, when administered flecainide (**Figure 4**). **Figure 4A** shows data from a Kir2.1^C122Y^ mouse with frequent closely coupled PVCs with compensatory pauses. In **Figure 4B**, data from a Kir2.1^G215D^ animals show several runs of monomorphic NSVT. **Figure 4C** is a recording of a 9:1 sinoatrial block pattern in a Kir2.1^Δ314–315^ mouse. Finally, **Figure 4D** shows an example with frequent episodes of NSVT of variable duration in a Kir2.1^R67W^ mouse.

**Figure 4.**
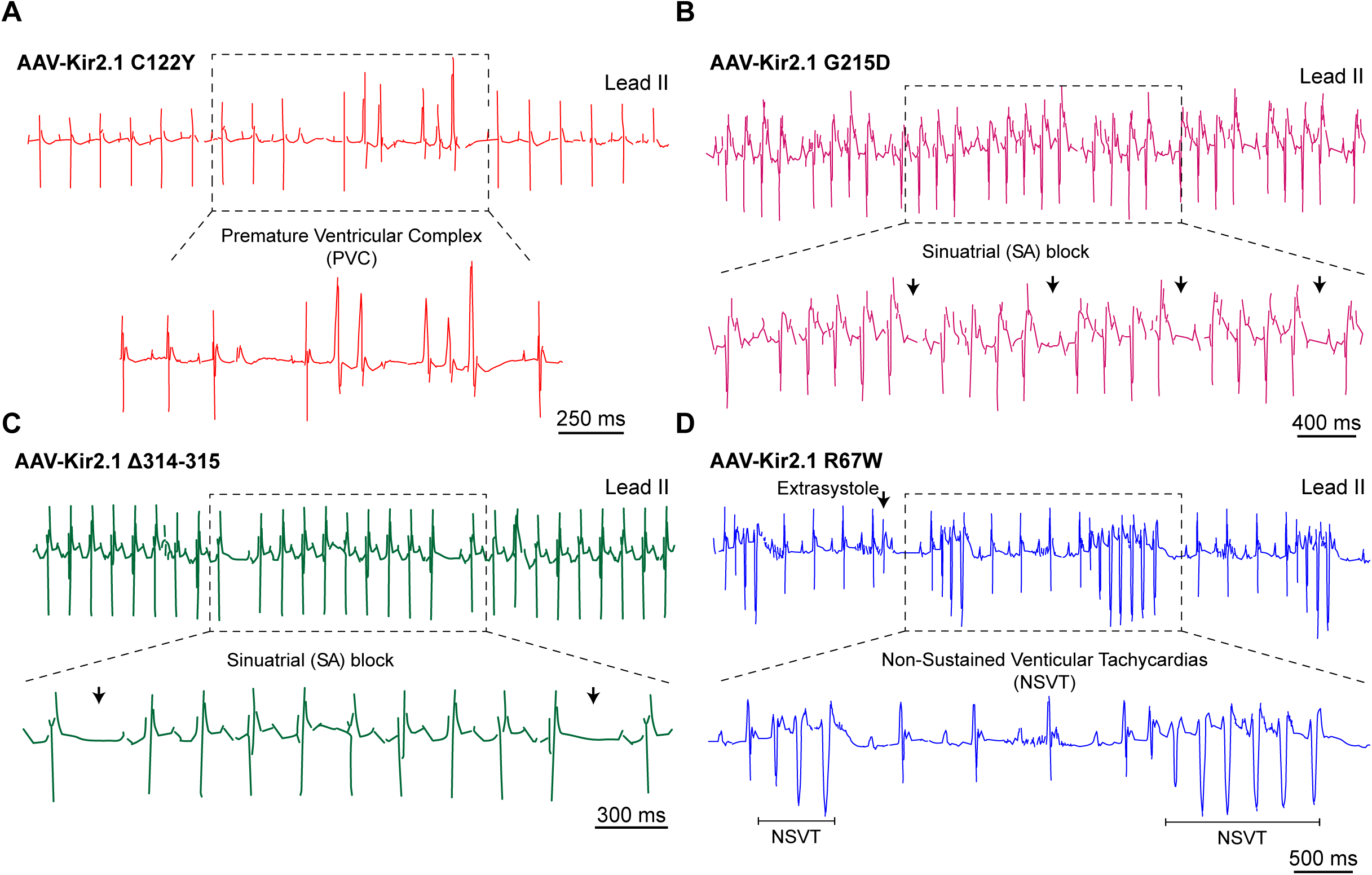
Flecainide is proarrhythmic and alters cardiac conduction in ATS1 mice. Representative electrocardiograms (ECG) lead-II traces recordings in AAV-transduced Kir2.1^C122Y^ (**A**) animals showing frequent premature ventricular complexes (PVCs), Kir2.1^G215D^ (**B**) and Kir2.1^Δ314–315^ (**C**) animals showing atrioventricular block, and Kir2.1^R67W^ (**D**) animals showing non-sustained ventricular tachycardia (NSVT) after flecainide administration.

### Flecainide fails to prevent arrhythmia inducibility in most ATS1 mutant mice

Our data compilation indicates that the cardiac electrical response of ATS1 patients to class-Ic AADs is highly variable. Propafenone has demonstrated limited success in managing VA compared to flecainide, and patients often require alternative treatment options. Therefore, hereafter we focus solely on flecainide effects on experimental animals and human cells. To test whether flecainide modifies arrhythmia susceptibility in ATS1 mouse models we conducted *in-vivo* catheter-based, intracardiac electrical stimulation experiments in all groups of animals (**Figure 5**). At baseline, all mutant animals (Kir2.1^Δ314–315^, 80%; Kir2.1^C122Y^, 50%; Kir2.1^G215D^, 50%, Kir2.1^R67W^, 66,67%; Kir2.1^S136F^, 62.5%) were inducible for VA developing NSVT episodes of variable duration. In contrast, only 1 out of 10 (10%) Kir2.1^WT^ mice was inducible (**Figure 5A and B, top**).

**Figure 5.**
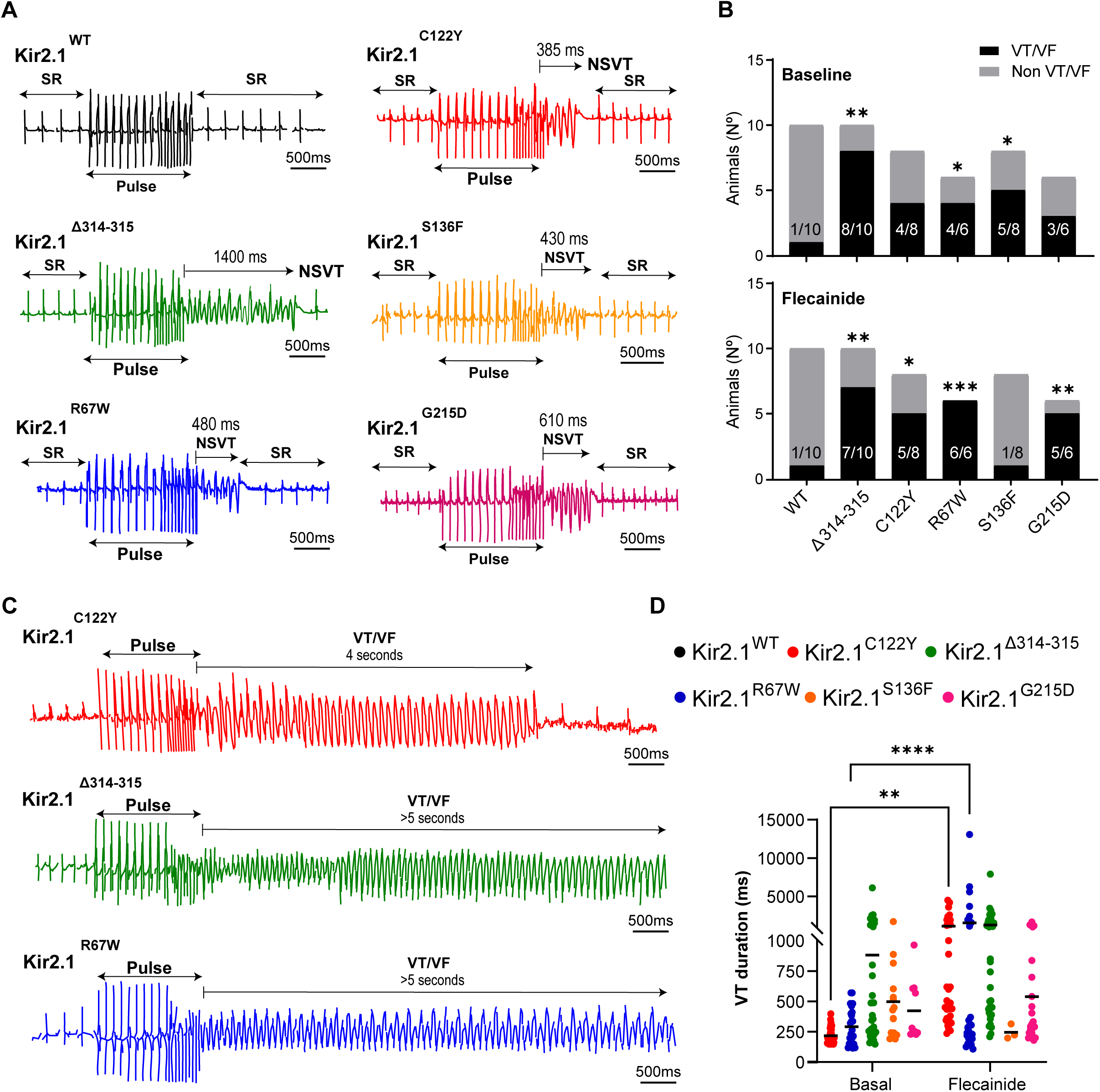
Flecainide increases susceptibility to arrhythmias in ATS1 mutant mice. **A**: Representative ECG lead-II traces after a train of intracardiac ventricular pulses in AAV-transduced Kir2.1^WT^ (black; N=10), Kir2.1^Δ314–315^ (green; N=8), Kir2.1^C122Y^ (red; N=8), Kir2.1^G215D^ (purple; N=6), Kir2.1^R67W^ (blue; N=6) and Kir2.1^S136F^ (orange; N=8) animals with periods of polymorphic ventricular tachycardia (PVT) at baseline. A short duration of non-sustained ventricular tachycardias (NSTV) is indicated. **B:** Contingency plots showing the number of animals with the arrhythmogenic response after intracardiac stimulation at baseline (top) and under flecainide (20mg/Kg) (bottom). **C**: Representative ECG lead-II trace showing longer NSVT runs after flecainide administration. **D**: Graph shows ventricular tachycardias (VT) duration at baseline and under flecainide administration. Statistical analysis using the Fisher’s exact test. * = p<0.05; ** = p<0.01; *** = p<0.001; **** = p<0.0001

Flecainide administration was beneficial in Kir2.1^S136F^ mice, reducing VT inducibility to 1 out of 8 animals (12.5%). However, a substantial increase in NSVT inducibility was demonstrated in the rest of mutant mice after a single dose of flecainide (40 mg/Kg; i.p.) (**Figure 5B bottom)**. In the presence of the drug, 70% of Kir2.1^Δ314–315^, 62.5% of Kir2.1^C122Y^, 83,34% of Kir2.1^G215D^; 100% of Kir2.1^R67W^; but only 10% of Kir2.1^WT^ mice were NSVT inducible. Importantly, although at baseline the mean duration of NSTV did not exceed 1 sec in any of the mutant mice, a single dose of 40 mg/Kg flecainide increased the duration of the induced VA to more than one second in all, except Kir2.1^G215D^ and Kir2.1^S136F^ mutant mice (**Figure 5C and D**). Such a variable response to flecainide resembles the heterogeneous clinical outcomes observed in ATS1 patients (**Table 1**).

We did observe significant alterations in sinus node function upon ventricular intracardiac stimulation in all mutant mice. We measured sinus node recovery time (SNRT) as the interval between the last pacing stimulus and the first atrial complex of sinus origin in recordings whose response to intracardiac stimulation was not arrhythmogenic (**Supplemental Figure 2**). At baseline, SNRT was longer in Kir2.1^Δ314–315^, Kir2.1^C122Y^ and Kir2.1^R67W^ mice than Kir2.1^WT^ (**Supplemental Figure 2A;** SNRT, 280 ± 98 ms in Kir2.1^Δ314–315^; 327 ± 65 ms in Kir2.1^C122Y^; 213 ± 69 ms in Kir2.1^G215D^; 291 ± 83 ms in Kir2.1^R67W^; and 196 ± 49 ms in Kir2.1^S136F^; *vs* 218 ± 48 ms in Kir2.1^WT^ mice). Notably, flecainide led to a significant SNRT increase in Kir2.1^Δ314–315^, Kir2.1^C122Y^ and Kir2.1^R67W^ mice, suggesting impaired sino-atrial excitability and conduction. In contrast, SNRT in Kir2.1^WT^, Kir2.1^G215D^ and Kir2.1^S136F^ mice showed no significant change compared to pre-treatment values (**Supplemental Figure 2B**; Kir2.1^Δ314–315^, 340 ± 116 ms; Kir2.1^C122Y^, 462 ± 182 ms; Kir2.1^G215D^, 240 ± 83 ms; Kir2.1^R67W^, 502 ± 192 ms; and Kir2.1^S136F^, 218 ± 76 ms ; *vs* Kir2.1^WT^, 224 ± 51 ms). While demonstrating therapeutic promise in Kir2.1^S136F^ animals, the results suggest that flecainide potentially harms sinus node function in Kir2.1^Δ314–315^, Kir2.1^C122Y^ and Kir2.1^R67W^ mice. In addition, the ECG also revealed prolonged P wave duration (**Figure 3**), suggesting delayed conduction through Bachmanńs bundle and the interatrial pathways, which is commonly associated with AF.^33^ Altogether, these results demonstrate a high variability in response to flecainide and suggest that susceptibility to proarrhythmia is dependent on the specific mutation. Thus, understanding the intricacies of flecainide’s actions in diverse genetic contexts is crucial for refining arrhythmia management strategies and improving ATS1 patient care.

### Flecainide impairs Kir2.1-Na_V_1.5 channelosome function in mutant cardiomyocytes

Flecainide and propafenone have been shown to increase I_K1_ by interfering with polyamine blockade in heterologous HEK293 cells expressing Kir2.1^WT^ channels,^34,35^ but this effect has not been demonstrated in adult cardiomyocytes. To gain insight into the ionic mechanisms of the proarrhythmic effects of flecainide in some of the ATS1 mice, we used equimolar therapeutic concentrations^21^ of the drug in patch-clamp experiments focusing on I_K1_ and I_Na_ (**Figure 6**). We abstained from analyzing isolated Kir2.1^Δ314–315^ cardiomyocytes due to the severe impact on I_K1_, which was completely absent.^20^ Superfusion of flecainide (10 μM) slightly increased I_K1_ in Kir2.1^WT^ and Kir2.1^S136F^ cardiomyocytes (**Figure 6A**). However, contrary to expected, this drug significantly reduced I_K1_ in Kir2.1^C122Y^ and Kir2.1^R67W^ cardiomyocytes. Flecainide did not modify I_K1_ in Kir2.1^G215D^ cardiomyocytes, as shown by the I/V plots and the I_K1_ slope values measured between -140 and -50 mV (**Figure 6A and C**). Notably, in Kir2.1^G215D^ cardiomyocytes, flecainide produced a small increase in the outward current, an effect that was nonexistent at baseline, although it did not exceed the levels observed in mutant Kir2.1^C122Y^ and Kir2.1^R67W^ cardiomyocytes (**Figure 6D**). On the other hand, compared to Kir2.1^WT^, peak I_Na_ density was significantly reduced only in Kir2.1^C122Y^ cardiomyocytes at baseline (**Figure 6B**). However, whereas 10 µM flecainide reduced I_Na_ density by ∼48% in Kir2.1^WT^, the drug further decreased it in Kir2.1^C122Y^ (∼62%), and Kir2.1^R67W^ (∼66%) cardiomyocytes, showing a weaker effect over Kir2.1^G215D^ (∼52%) and Kir2.1^S136F^ (∼53%) cells (**Figure 6E-F**). These data further demonstrate that ATS1 mutants differentially modify I_K1_ and I_Na_ densities at baseline and under flecainide administration. Thus, our results reinforce the hypothesis that conduction disturbances and arrhythmias in ATS1 patients are not only due to I_K1_ and RMP defects but also reduced excitability exacerbated by flecainide.

**Figure 6.**
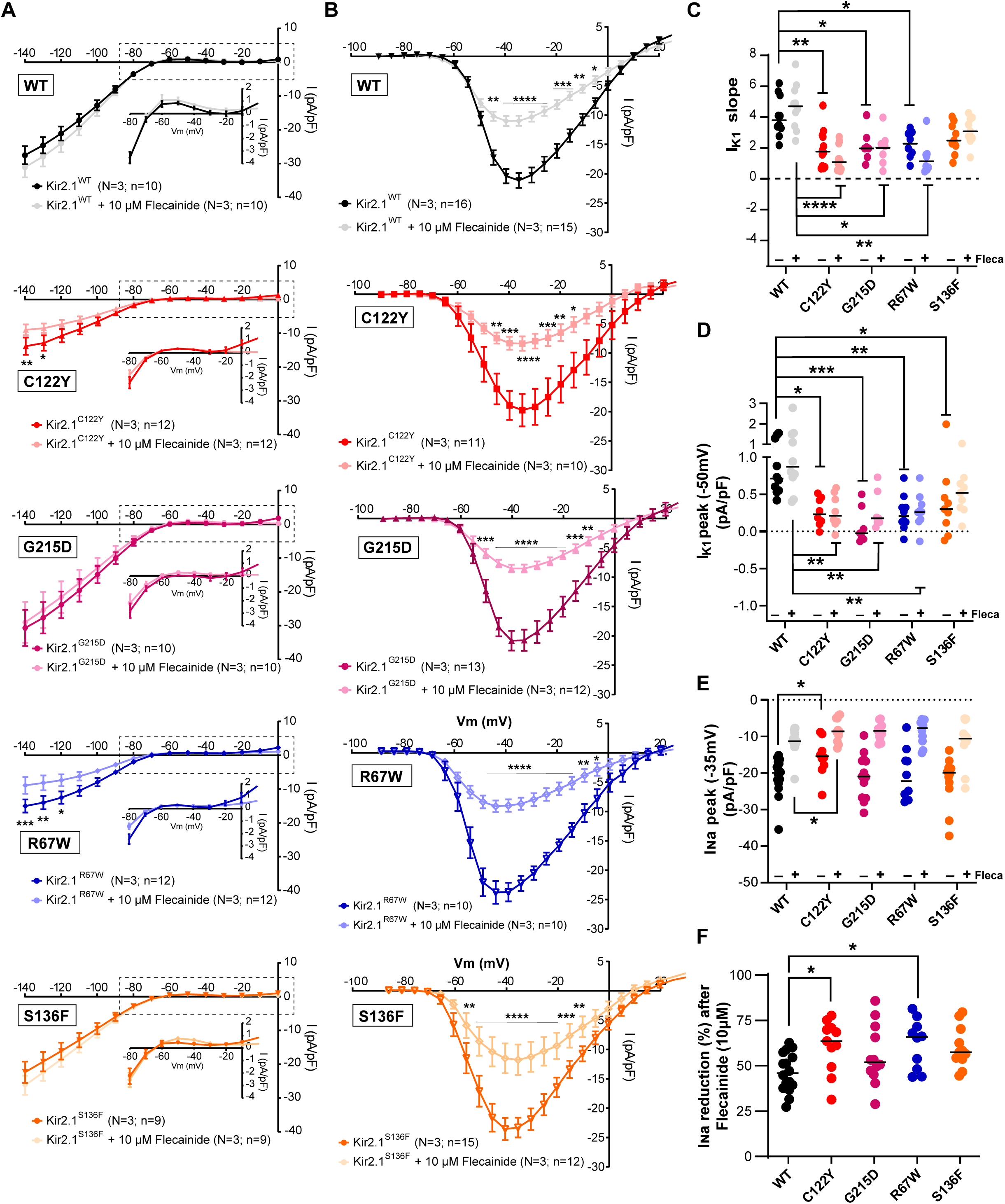
Cardiac expression of ATS1 mutations alter mouse ventricle electrophysiology in isolated cardiomyocytes. **A**: Current-voltage (I/V) relationships of inward rectifying potassium current I_K1_ and **B**: I_Na_ density in Kir2.1^WT^ (black), Kir2.1^Δ314–315^ (green), Kir2.1^C122Y^ (red), Kir2.1^G215D^ (purple), Kir2.1^R67W^ (blue) and Kir2.1^S136F^ (orange) cardiomyocytes at baseline and under flecainide superfusion. **C**: I_K1_ slope from -140 to -60 mV at baseline and under flecainide administration. **D**: Sodium density I_Na_ peak (pA/pF) at -35mV. **E**: Percentage of remaining I_Na_ density after flecainide inhibition. Statistical analyses were conducted using two-tailed ANOVA. * = p<0.05; ** = p<0.01; **** = p<0.0001.

### ATS1 patient-specific iPSC-CMs are highly inducible for re-entrant arrhythmias

We generated hiPSC-CM monolayers from consenting ATS1 patients carrying Kir2.1^C122Y^, Kir2.1^R67W^ and Kir2.1^G215D^ mutations (see *Supplementary Methods*). We conducted optical mapping experiments to validate our previous experiments and better understand the variable electrophysiological phenotypes of the patients in response to flecainide. Importantly, we used a CRISPR-mediated genetically corrected isogenic control (IC) for all three patient-specific hiPSC-CM to fully assess the contribution of Kir2.1 mutations to arrhythmia. We carried out optical mapping experiments using the voltage-sensitive fluorescent dye FluoVolt in monolayers paced at basic cycle lengths (BCL) between 400 and 1000 ms. To better understand the underlying mechanism(s) of flecainide-induced arrhythmogenesis, mutant and IC monolayers were superfused with 5 µM flecainide. CV restitution curves constructed from color phase maps displayed slower velocities in C122Y iPSC-CMs monolayers compared to IC. Impressively, the CV was highly reduced in C122Y monolayers after flecainide superfusion (**Figure 7B**). Additionally, flecainide induced non-sustained re-entrant activity with complete rotations followed by spontaneous termination in patient-specific Kir2.1^C122Y^, Kir2.1^R67W^ and Kir2.1^G215D^ iPSC-CMs **(Figure 7C)**. Mutant iPSC-CMs monolayers were more inducible for re-entrant arrhythmias compared to ICs (6 out of 11, 54.5% in C122Y iPSC-CMs *vs* 2 out of 11, 18.2% in IC; 6 out of 11, 54.5% in R67W *vs* 3 out 9, 33.3% in IC; 7 out of 11, 63.6% in G215D *vs* 5 out of 11, 45.5% in IC) (**Figure 7D**). Most importantly, flecainide superfusion resulted in an increased rate of rotor generation in C122Y, and R67W monolayers (rotors; 8 out of 11, 72.7% in C122Y *vs* 4 out of 14, 28.6% in IC; 8 out of 11, 72.7% in R67W *vs* 2 out of 6, 33.3% in IC). However, while flecainide eliminated arrhythmias in IC monolayers (5 out of 11, or 45,5% rotors in untreated G215D IC *vs* 3 out of 10, or 30% rotors in flecainide-treated G215D IC), it did not alter arrhythmia inducibility in G215D iPSC-CMs; i.e., rotors were maintained during flecainide superfusion (7 out of 11, 63.6% in non-treated G215D *vs* 7 out of 11, 63.6% in flecainide-treated cultures) (**Figure 7D**). Taken together, these data demonstrate that flecainide exacerbates the already abnormal conduction in patient-specific iPSC-CMs^C122Y^ monolayers, leading to reentry and polymorphic tachycardia. However, the molecular mechanisms underlying arrhythmia inducibility and the ineffectiveness of flecainide in R67W and G215D iPSC-CM monolayers, respectively, remain unclear and suggest more complex mechanisms that are beyond the scope of this study.

**Figure 7.**
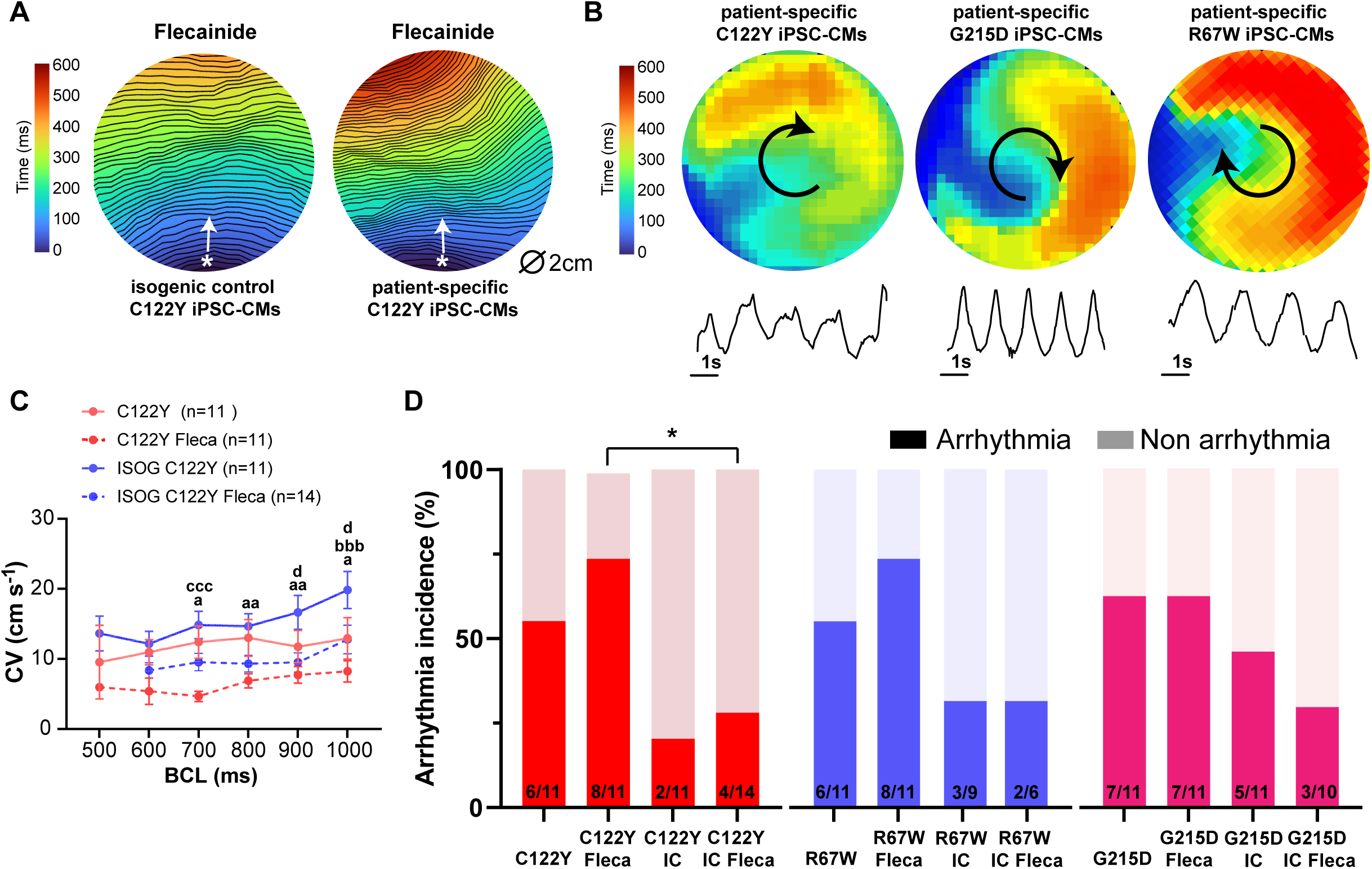
Flecainide leads to conduction defects and re-entrant arrhythmias in patient-specific iPSC-CMs. **A:** Representative single camera pixel recording from an optical mapping experiment show velocity maps with 1 ms activation isochrones in patient-specific iPSC-CMs^C122Y^ (red) and CRISPR-mediated isogenic control iPSC-CMs^C122Y^ (blue) hearts paced at a basic cycle length (BCL) of 400 ms. The color bar indicates conduction velocity (CV; cm s-1). The white asterisks indicate the pacing point and the propagation direction is indicated by the white arrow. **B**: Re-entrant arrhythmias from C122Y, G215D and R67W patient-specific iPSC-CMs monolayers under flecainide treatment. Below each map is a single pixel recording revealing different patterns of monomorphic or polymorphic re-entrant tachycardia maintained by one self-sustaining rotor. **C**: The CV restitution curve displayed slower velocities in iPSC-CMs^C122Y^ monolayers at all frequencies tested. All groups presented slightly slower velocities at higher frequencies. Each value is the mean ± SEM (N=5 differentiations; n indicates number of monolayers per condition; two-way ANOVA corrected by Tukeýs multiple comparisons test, * p<0.05; ** p<0.01; a=IC C122Y vs C122Y; b= IC C122Y Fleca vs C122Y Fleca; c= IC C122Y vs 122Y Fleca; and d= IC 122Y vs IC C122Y Fleca). **D**: Contingency plots of number of monolayers show arrhythmia inducibility for each group. Data show a high rate of arrhythmia susceptibility in mutant iPSC-CMs. Each value is the mean ± SEM (Fisher’s exact test for contingency data).

### ATS1 mutations differentially alter flecainide-to-Kir2.1 molecular docking

Previous computational studies have predicted that flecainide and propafenone share a common binding site within the tetrameric Kir2.1 channel determined by a pharmacophore involving Cys_311_ in the cytosolic space.^34^ However, it has not been determined whether Kir2.1 mutations may differentially reorganize the global structure of the Kir2.1 channel and destabilize the pharmacophore binding site determined by Cys_311_, interfering with flecainide binding. In line with this, recent findings show that the extracellular Cys_122_-to-Cys_154_ disulphide bond breaks disrupt Kir2.1-PIP_2_ interaction in the cytosolic-transmembrane region, which is distant from the location of the Kir2.1^C122Y^ mutation.^14^ To test the above hypothesis, we used *in-silico* molecular docking modeling to derive predictions of the molecular interaction of the mutated Kir2.1^C122Y^ channel and flecainide in the Cys_311_-mediated pharmacophore.

Our simulations revealed that Kir2.1^WT^ channels exhibit a substantially enhanced binding affinity to flecainide in the presence of PIP_2_, which is crucial for stabilizing in the open state the narrowest part of the ion conduction pore (G-loop)^36,37^. Defects in PIP_2_ binding are a major pathophysiological mechanism underlying the loss-of-function phenotype for several ATS1-associated mutations.^9,38–40^ The atomic binding sites for flecainide and PIP_2_ are closely located, and our simulation suggests that PIP_2_ could help accommodate flecainide molecules. Accordingly, in the absence of PIP_2_ flecainide failed to bind to Cys_311_ in Kir2.1^WT^ chains **c** and **d** in our simulations (**Figure 8A)**. Impressively, a complete set of flecainide molecules (one per Kir2.1^WT^ monomer) effectively integrated in the presence of PIP_2_, exhibiting enhanced binding energy compared to its absence (**Figure 8B)**. This was evidenced by the Gibbs free-energy interface of flecainide binding in the presence of PIP_2_ in chains **a** (ΔdG-11.388) and **ab** (ΔdG-12.868 for chain **a** and ΔdG-13.278 for chain b) *vs* chains **a** (ΔdG-9.948) and **ab** (ΔdG-9.979 for chain **a** and ΔdG-9,102 for chain **b**) in absence of PIP_2_ (**Figure 8A-B**). Significantly, even though the Kir2.1^C122Y^ mutation alters the PIP_2_ binding pocket,^14^ we observed that flecainide was fully incorporated into the heterozygous mutant channels with a total of 2 PIP_2_ molecules (one per Kir2.1^WT^ monomer) (**Figure 8C**).

**Figure 8.**
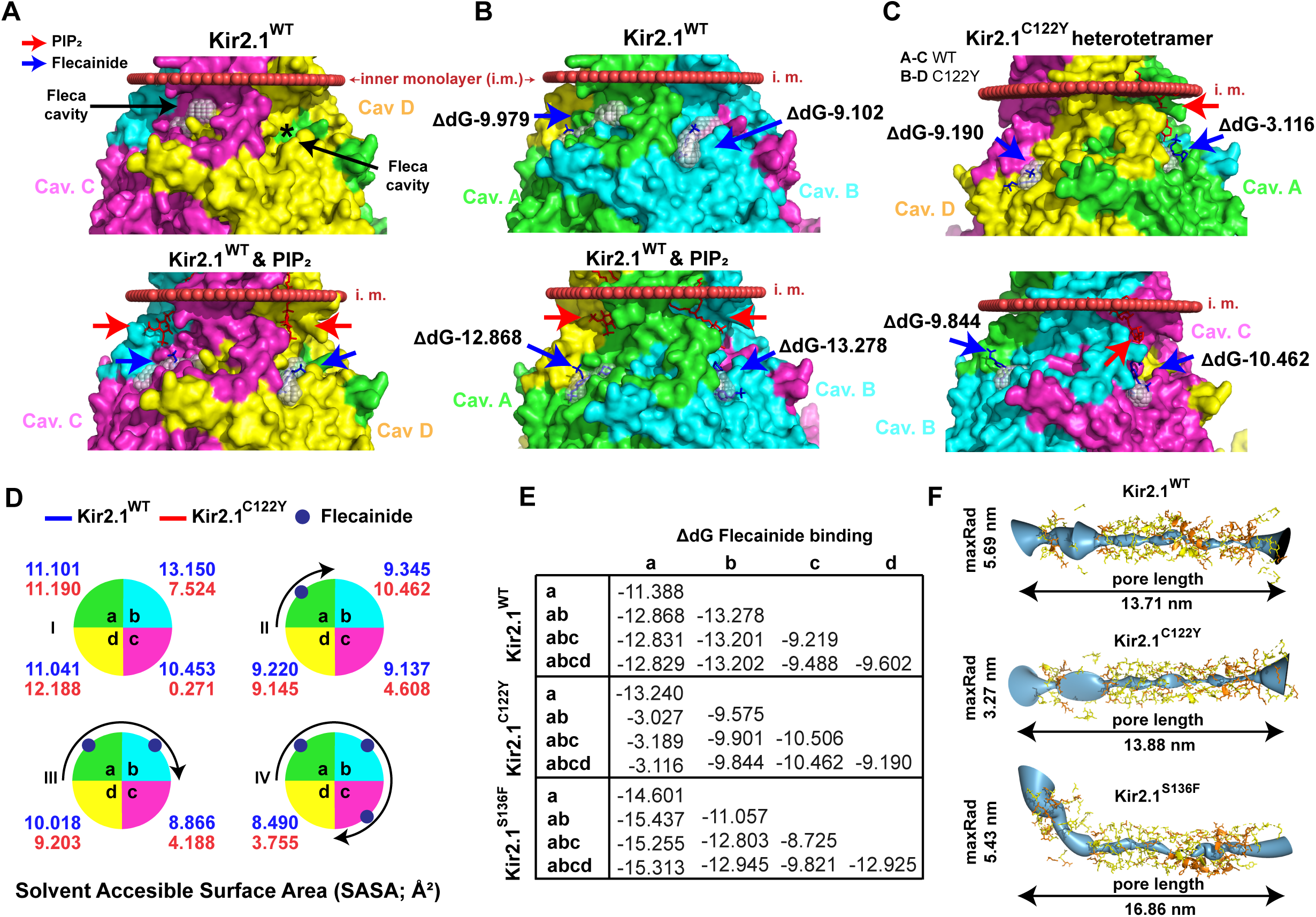
Kir2.1 mutated channels display conformational alteration in the pharmacophore binding site at Cys_311_. **A:** Tridimensional (3D) representation of Kir2.1^WT^ showing cavities C (magenta) and D (yellow) in the absence (top) and presence (bottom) of PIP_2_ molecules. Note that PIP_2_ is required for proper flecainide binding in **c** and **d** cavities. PIP_2_ (red arrow) and flecainide (blue arrow) are also indicated. **B:** 3D representation of flecainide binding in cavity **a** (green) and **b** (blue) in the absence (top) and presence (bottom) of PIP_2_ molecules in Kir2.1^WT^. Gibbs free energy values are shown, being more stable in the presence of PIP_2_. **C:** 3D representation of flecainide binding in Kir2.1^C122Y^ heterotetramer. Flecainide is fully present in Kir2.1^C122Y^ heterotetramer. **D:** Solvent Accessible Surface Area (SASA) of each Cys_311_-pharmacophore in Kir2.1^WT^ (blue) and Kir2.1^C122Y^ (red) in sequential incorporation flecainide molecules. **E:** Comparative table of Gibbs free energy values of flecainide binding to each chain in Kir2.1^WT^, Kir2.1^C122Y^ and Kir2.1^S136F^ channels. **F:** Schematic representation of Kir2.1^WT^ (top), Kir2.1^C122Y^ (middle) and Kir2.1^S136F^ (bottom) channel showing a similar pore length (13.71 nm; 13.88 nm; and 16.86 nm) and maxRadius (5.69 nm; 3.27 nm; and 5.43 nm), respectively.

However, the Solvent Accessible Surface Area (SASA) of each Cys_311_-pharmacophore in mutant C122Y heterotetramers exhibited significant variation along the contiguous chain as we introduced flecainide molecules, indicating a substantial decrease in the exposed surface area, which could potentially disrupt stable flecainide binding (**Figure 8D**). The incorporation of a second flecainide molecule in the mutant heterotetramer dramatically decreased the Gibs free-energy of the firstly incorporated flecainide, as indicated the values for chains **a** (dG -13.24) and **ab** (ΔdG -3.027 for chain **a** and ΔdG -9.575 for chain **b**) (**Figure 8E**). Comparative analysis of channel properties revealed a similar pore length in WT (13.71 nm) and mutant (13.88 nm) heterotetramers, but with more than 40% reduction in maxRadius (5.69 nm in WT vs. 3.27 nm in mutant heterotetramer), suggesting channel constriction in the presence of flecainide (**Figure 8E**). In contrast, simulation of the Kir2.1^S136F^ mutant heterotetramer did not disrupt the Cys_311_ binding pocket, and flecainide successfully incorporated in mutant heterotetramers showing similar channel properties and binding capacity to the Kir2.1^WT^ model (**Figure 8E-F**). Incorporation of a second flecainide molecule retained the Gibs free-energy of the firstly incorporated flecainide for chains **a** (ΔdG -14.601) and **ab** (ΔdG -15.437 for chain **a** and ΔdG -11.057 for chain **b**) (**Figure 8E**). Analysis of channel properties revealed a similar maxRadius (5.43 nm) and an increased pore length (16.86 nm) compared to the Kir2.1^WT^ structure (**Figure 8F**).

Collectively, these *in silico* homology studies predict that the Cys_311_ binding pocket is not conserved in mutant Kir2.1 channels. Depending on the specific mutation, the Cys_311_ binding pocket may undergo unique atomic rearrangements that affect both the accessibility and energetic stability of flecainide molecules. This could potentially disrupt the dynamics of flecainide binding, leading to different outcomes for channel function depending on the particular ATS1 mutation.

## Discussion

Flecainide received Food and Drug Administration (FDA) approval in 1984 for the treatment of sustained VT. Subsequently, the Cardiac Arrhythmia Suppression Trial (CAST) found that the drug was proarrhythmic and associated with excess all-cause mortality in patients with structural heart disease.^41^ Since then, class-Ic AADs are contraindicated in patients with coronary heart disease or chronic congestive heart failure. Class-Ic AADs are potent Na^+^ channel blockers that produce profound reductions in conduction velocity (CV)^42,43^ and facilitate reentry, which explains their proarrhythmic effect.^44^ These agents do not often suppress sustained VA induction by programmed stimulation.^45,46^ They raise the defibrillation threshold in experimental models,^47,48^ and their use has been associated with incessant VT resistant to cardioversion.^49,50^ On the other hand, both flecainide and propafenone have been indicated for supraventricular arrhythmias, including atrial fibrillation (AF). To the best of our knowledge, the mechanism has not been elucidated. Paradoxically, some investigators attribute such effects to chamber-specific differences with greater flecainide inhibition of atrial I_Na_, leading to higher reduction of both maximum AP upstroke velocity and CV in atrial tissue.^51^ Such an interpretation seems counter to the well-established knowledge that reduced maximum AP upstroke velocity and slowed conduction are pro-arrhythmic. Clinical and molecular evidence suggest that flecainide could potentially promote arrhythmias even in patients with structurally normal hearts, including reports of TdP and life-threatening PVT without coronary artery disease or heart failure.^52,53^ Despite these risks, class-Ic AADs have been successfully used to treat hundreds of patients, and clinical guidelines continue to recommend them for specific arrhythmogenic cardiac diseases.^7,8,54,55^

ATS1 is a rare disease that may lead to life-threatening VA and SCD without affecting heart structure^3^. More than 90 ATS1-associated autosomal dominant mutations have been identified in the Ki2.1 protein. However, the molecular mechanisms are not fully understood, and variability in phenotype severity, even within the same family^31^, complicates treatment. Current antiarrhythmic therapies include β-blockers and class-Ic AADs. The use of β-blockers has been derived from an early view that considered ATS1 as a variant of LQTS^1^, and from the assumption that patients would respond to antiadrenergic interventions, despite the lack of robust mechanistic underpinnings. In addition, a common treatment for ATS1 is flecainide or propafenone alone or in combination with β-blockers.^56–59^ The use of class-Ic AADs in patients with ATS1 started with a report suggesting that flecainide effectively suppressed isolated PVCs and NSVT.^60^ Since then clinical guidelines recommend combinations of β-blockers and traditional AADs like flecainide and propafenone.^7,8^ However, their efficacy and safety in treating ATS1 remain contentious, and emerging evidence suggests that they may be associated with variable efficacy and potential proarrhythmic effects.

To date, clinical guidelines fail to encourage standardization and homogeneity in defining VA and therapeutic success, as these vary significantly among reports, representing a mayor limitation of the available data. In support of this clinical observation, in our analysis of 53 ATS1 reported patients, flecainide and propafenone showed partial effectiveness. As illustrated in **Table 1**, several brief reports have suggested that flecainide, alone or in combination with β-blockers suppresses VA in ATS1 patients.^4,57,58,60–72^ One study reported a 54-year-old male carrying the Kir2.1^302M^ mutation who experienced over 98% reduction in the number of PVCs (from a total of 15671 in 24h Holter monitoring) following flecainide treatment (200mg/day).^62^ However, a careful review of clinical data from 48 patients showed a large patient-to-patient variability, mostly with incomplete disappearance of VA under flecainide, which did not prevent life-threatening arrhythmias and SCD (**Table 1**).^4,57,62,64,73^ In a case report of a 15-year-old boy carrying the Kir2.1^R218W^ mutation, flecainide, mexiletine or nadolol monotherapy was ineffective in preventing or reducing VA; only a combination of flecainide with verapamil reduced the number of PVCs by about 87% (from 40846 per day).^64^ In contrast, patients experienced more moderate responses to flecainide alone. For example, ATS1 probands with the Kir2.1^R67Q^ mutation showed only about 44% VA reduction (from a total of 19389 to 10854 PVCs) under flecainide (100 mg/day) combined with atenolol (50mg/day).^73^ Additionally, Miyamoto et al. reported that oral flecainide (100-200 mg/day) reduced VA in 10 patients, who still maintained a ∼30% VA burden.^41^ Several reports have shown that class-Ic AADs like flecainide and propafenone are ineffective in suppressing VA in ATS1.^4,14,74–78^ In one case, propafenone was ineffective even when combined with verapamil, as the proband still presented frequent episodes of bigeminy^75^. Importantly, some studies have reported cases of aborted SCD in patients with ATS1 who received an external defibrillation or an ICD shock under treatment with flecainide or propafenone.^4,14,60,72,74,77^ Although flecainide was effective in reducing the high burden of VA in one patient, Bokenkamp et al. reported one appropriate ICD shock for Torsades de Pointes (TdP) tachycardia after three years under flecainide. In contrast, the patient’s sister was asymptomatic carrying the same Kir2.1^R218W^ mutation.^60^ In the series of Delannoy et al, one patient experienced a non-fatal cardiac arrest related to NSVT despite β-blockers and flecainide treatment.^77^ Similarly, Mazzanti et al. reported that the combination of these drugs usually fails to reduce arrhythmias in ATS1.^4^ Interestingly, in Mazzantís series 2 out of 12 monitored probands experienced non-fatal cardiac arrest while on treatment^4^, but it remains unclear whether this was due to proarrhythmia or merely ineffective therapy.

In our analysis, more than 20 loss-of-function Kir2.1 mutations were associated with highly variable phenotypes (**Table 1**). This suggests that the molecular mechanisms underlying the increased susceptibility to arrhythmias and SCD in ATS1 patients may differ based on the specific mutation, which is crucial in determining the response to AADs. In addition, our animal and human cell experiments reinforce the idea that ion channel interactions within multiprotein complexes, including Kir2.1-Na_V_1.5 channelosomes, likely contribute to the wide variety of clinical outcomes.^31,79–81^ Therefore, the molecular and functional interactions between Kir2.1-Na_V_1.5 indicates that ATS1 should no longer be considered in simplistic terms as a “monogenic” disorder. Reduction in I_K1_ may directly affect the functional expression or biophysical properties of Na_V_1.5 channels. I_K1_ reduction depolarizes the RMP and reduces the availability of Na_V_1.5 for activation, with consequent impairment of cellular excitability and CV.^82–84^ However, recent studies have shown that some mutation can also directly impact I_Na_ density, adding another layer of complexity to the electrophysiological alterations in ATS1. For instance, the trafficking-deficient Kir2.1^Δ314–315^ mutation and the Kir2.1^C122Y^ mutant that affects Kir2.1-PIP_2_ interactions reduce Kir2.1 and Na_V_1.5 protein levels at the cell membrane interfering with both I_K1_ and I_Na_ densities in the cardiomyocyte. Both mutations lead to a slow-conduction substrate favorable for cardiac arrhythmias.^23,14^ The balance between I_Na_ and I_K1_ is a key determinant of the frequency and stability of ventricular rotors that ultimately generate cardiac fibrillation and lead to SCD.^85^ This delicate balance could be disrupted by some ATS1 mutations at baseline and further exacerbated by class-Ic AADs. Therefore, pharmacological treatment and clinical management should be carefully tailored based on the specific mutation and individualized for each patient. However, to our knowledge, no previous studies have evaluated the proarrhythmogenic consequences of ATS1 and their impact on cardiac electrophysiology in response to class-Ic AADs.

Our finding revealed that flecainide and propafenone differentially prolonged the P wave duration and the PR, QRS and QTc intervals. They also increased VA inducibility compared with Kir2.1^WT^. In addition, flecainide promoted spontaneous VA and conduction disturbances of varying types (**Figure 4**), including NSVT, atrioventricular (AV) block, and long-short coupling PVCs, which have been previously associated with Purkinje fiber dysfunction.^86^ Altogether, these observations support the presence of different mutant-dependent mechanism, consistent with our hypothesis. Further, voltage-clamp experiments revealed that flecainide increased I_K1_ density in Kir2.1^WT^ cardiomyocytes, as previously described in HEK293 cells.^34,35^ However, in support of our hypothesis, the effects of flecainide were different in cardiomyocytes expressing mutant channels (**Figure 6)**. While Kir2.1^S136F^ responded similar to Kir2.1^WT^, flecainide resulted ineffective in recovering the already decreased I_K1_ of Kir2.1^G215D^ cardiomyocytes. Interestingly, in both Kir2.1^R67W^ and Kir2.1^C122Y^, I_K1_ density was significantly reduced both under basal conditions and in the presence of flecainide, which emphasizes I_K1_ blockade as a pro-arrhythmic mechanism of flecainide and a possible mechanism for prolonged QTc. Similarly, the class III AAD amiodarone blocks IK1 ^87^ and likely reduces excitability by depolarizing the RMP giving a rationale for an increased risk of life-threatening arrhythmic events in ATS1 patients. As Mazzanti et al also suggested, amiodarone may be pro-arrhythmic and should be avoided in ATS1 patients.^4^ Flecainide also exacerbated the reduction of I_Na_ density despite normal channel trafficking in Kir2.1^C122Y^ and Kir2.1^R67W^, and with great dispersion in Kir2.1^G215D^ mutant cardiomyocytes, which suggests heterogeneity that contributes to proarrhythmia. Accordingly, in ATS1 patient-specific iPSC-CM monolayers flecainide reduced CV in C122Y monolayers. Nevertheless, based on the above findings, flecainide was ineffective in modifying the high rotor incidence in G215D cultures compared to baseline but increased in C122Y and R67W, supporting the druǵs proarrhythmic effect.

Flecainide was originally characterized as a potent Na^+^ channel blocker that produce profound reductions in conduction velocity (CV),^42,43^, but flecainide can also affect ionic currents governing ventricular repolarization,^88–91^ although the molecular mechanisms are poorly understood. Clinical observations support the notion that cardiac electrical instability during flecainide treatment may be determined by defects in ventricular repolarization.^92–95^ Flecainide blocks the transient outward K^+^ current (I_to_) in a concentration-dependent manner, and some experimental studies attribute its proarrhythmic effect in structurally normal hearts to increased spatial repolarization gradients^96^ brought about by varying I_to_ expression across the myocardial wall.^97^ But, flecainide can also induce arrhythmia by increasing repolarization heterogeneities at the level of the Purkinje fiber-ventricular muscle junction^98^. Non-uniform expression of the delayed rectifier K^+^ channels throughout the ventricular epicardium may also contribute to dissimilar flecainide-induced APD lengthening in the left *vs* right ventricles.^99,100^ In addition, flecainide modulates ryanodine receptor type 2 (RyR2) activity and inhibits the RyR2 open state to prevent diastolic Ca^2+^ waves that trigger arrhythmias.^7,101,102^. Flecainide might also modify the recently demonstrated SR microdomain of functional Kir2.1 channels^103^ that provide countercurrent to SERCA-mediated Ca2^+^ influx, potentially altering Ca^2+^ handling, which may contribute to proarrhythmia in ATS1.^23^ More studies would be needed to assess the mutant-dependent contribution on Ca^2+^ homeostasis and VA in flecainide-treated ATS1 patients.

While flecainide and propafenone interact with Cys_311_ in the Kir2.1^WT^ subunits and reduce polyamine-induced inward rectification, it remains unclear how mutant ATS1 channels impact the Cys_311_ pharmacophore. Our in-silico molecular docking experiments indicate that, unlike Kir2.1^WT^ channels, the Cys_311_ binding pocket is not conserved in ATS1 mutant heterotetramers. This might disrupt the dynamic maintenance of flecainide binding with unexpected consequences, potentially leading to channel closure and arrhythmias in a subset of mutant channels, as we observed *in-vivo*. Interestingly, our in-silico studies also predict that the Kir2.1-PIP_2_ interaction energetically stabilizes the binding of flecainide to the Kir2.1 channel. These findings suggest that mutations like C122Y, R67W, and G215D that disrupt Kir2.1-PIP_2_ interactions^9,14^ could result in less favorable outcomes compared to mutations like S136F, which do not interfere with this interaction but affect filter-selectivity properties.^9,31,32^ However, further studies are required to elucidate whether mutation-induced disruption of Kir2.1-PIP_2_ interactions is a key factor in the pro-arrhythmogenic effects of flecainide, which would lead to its contraindication in patients carrying such mutations. Studies should also explore the molecular effects of incorporating others AADs alone or in combination with β-adrenergic blocking drugs.

The results of our study underscore the need for a cautious re-evaluation of class-Ic AADs in ATS1 patients to achieve optimal treatment outcomes and minimize adverse effects. Our results suggest that Kir2.1 mutations should not only be classified according to their location in the channel structure but also based on their functional implications at the subcellular and cellular levels. In this sense, cardiac excitability is likely to be dramatically reduced in ATS1 patients carrying the trafficking-deficient Kir2.1^Δ314–315^ mutation, for whom we suggest direct exclusion of sodium channel blockers like flecainide and propafenone, because of the high risk of arrhythmias. The same would be expected for the Kir2.1^C122Y^ mutation which, despite reaching the sarcolemma, its structural changes lead to its Kir2.1-Na_V_1.5 channelosome degradation.^14^ Conversely, patients carrying mutations like Kir2.1^S136F^, which does not reduce excitability and CV, might benefit from the use of currently available AADs as demonstrated. In other words, a specific combination of β-blockers and flecainide might be appropriate for ATS1-causing mutations that alter the conformation of the channel, but not for mutations that affect Kir2.1 trafficking or others that lead to a decrease in I_Na_ density.

### Limitations

Like any other rare disease, implementing evidence-based therapeutic interventions for ATS1 presents significant challenges. Conducting conventional randomized controlled trials on ATS1 is particularly difficult due to the minimal number of patients worldwide. Unfortunately, the reported data often lack uniformity in key details, such as treatment duration, dosages and routes of administration. As such, after exhaustively searching the literature, we summarized data from only 23 publications containing isolated ATS1 case reports or small series without follow-up. Our study is limited to assessing the effects of an empirical, “one-size-fits-all” antiarrhythmic treatment of patients of both genders, all age ranges and diverse origins based on flecainide and other class-Ic AADs. Such limitations notwithstanding, we demonstrate that the cardiac electrical response of ATS1 patients to class-Ic AADs is highly variable, and at times contrary to anticipated. While some patients benefit from the use of these drugs, others do not, and yet others may get worse. The results support our hypothesis that the effect of class-Ic AAD treatment in a given ATS1 patient will depend on the precise electrophysiological consequences of the Kir2.1 loss-of-function mutation. The hypothesis is further supported by experiments in a small but highly relevant group of mutant mouse models, iPSC-CMs from three different ATS1 patients and in-silico simulations demonstrating a significant hierarchy in the mutation-induced arrhythmic phenotype and molecular effects in response to flecainide treatment. While well outside of the scope of this study, we are well aware of the need to assess the role of channels other than Kir2.1 and Na_V_1.5 and the effects of spatial heterogeneities to establish more rigorously the mechanisms of mutation-induced arrhythmias and their exacerbation by the drugs. Nevertheless, our experimental results strongly support that flecainide-induced pro-arrhythmia might be at least in part due to specific reductions in both I_K1_ and I_Na_, which significantly impairs excitability as demonstrated by the prolongation of the PR and QRS intervals in the mutant mice and the slowing of CV in the mutant iPSC-CMs.

### Conclusion

The results presented here open new horizons for the pharmacologic management of ATS1 patients. They demonstrate for the first time that whether or not class-Ic AADs will benefit a given ATS1 patient will depend on the precise electrophysiological consequences of specific Kir2.1 mutation. As such, we must no longer correct only the lack of function of the mutated channel, but also consider the possible repercussions on the molecular interactors of the channel that each particular mutation entails. A deeper understanding of the different arrhythmogenic mechanisms associated with different ATS1 mutations should open new pathways for personalized treatments of patients suffering from these devastating channelopathies. Our results emphasize the need to gain a deeper understanding of the specific pharmacologic regulation of Kir2.1 mutations, elucidate the pathophysiology of arrhythmias in individual patients with ATS1, and identify an effective, more personalized therapy that reduces individual proarrhythmia risk. Therefore, studies on the usefulness of already approved drugs and their derivatives for the treatment of diseases such as ATS1 should continue to improve the stratification of affected patients.

## Data Availability

The authors declare that all supporting data are available within the article.

## Non-standard Abbreviation and Acronyms

AAD: Antiarrhythmic drug
AP: Action potential
APD: Action potential duration
ATS1: Andersen-Tawil Syndrome Type 1
BiVT: Bidirectional ventricular tachycardia CV Conduction velocity
ECG: Electrocardiogram
hiPSC-CMs: Induced pluripotent stem cell-derived cardiomyocytes
IC: Isogenic control
ICD: Implantable cardiac defibrillator
i.p.: Intraperitoneal
i.v.: Intravenously
NSVT: Nonsustained ventricular tachycardia
PIP_2_: Phosphatidylinositol-4, 5-bisphosphate
PV: Premature ventricular contraction
PVT: Polymorphic ventricular tachycardia
RMP: Resting membrane potential
SASA: Solvent Accessible
SCD: Sudden cardiac death
SNRT: Sinus node recovery time
TdP: Torsades de pointe
VA: Ventricular arrhythmias
VE: Ventricular extrasystoles
VT: Ventricular tachycardia
WT: Wild type

## ACKNOWLEDGEMENTS

We thank the CNIC Viral Vectors Unit for producing the AAV9. Confocal experiments were conducted at the CNIC Microscopy and Dynamic Imaging Unit. We thank the CNIC Bioinformatics Unit for generating the *in-silico* homology modeling simulations, F-function analysis, and helpful discussions. We also thank the Centro de Supercomputación de Galicia (CESGA) for the use of the Finis Terrae III supercomputer to perform molecular dynamics studies. The CNIC is supported by the Instituto de Salud Carlos III (ISCIII), the Ministerio de Ciencia, Innovación y Universidades (MICIU), and the Pro CNIC Foundation, and is a Severo Ochoa Center of Excellence (grant CEX2020-001041-S funded by MICIN/AEI/10.13039/501100011033).

## AUTHOR CONTRIBUTION

F.M.C. and J.J. co-designed the experiments; F.M.C. performed most of the experiments; A.I.M.M. and P.G.S. are authors for cellular electrophysiology; E.Z., and J.J.J. provided clinical data, discussion and revisions; F.M. were in charge of in-silico homology modeling and molecular docking studies; P.S.P., M.L.V. and A.T.G. cultivated iPSC-CMs cells; L.K.G. and J.R.R performed optical mapping analysis; A.M., G.M.P., S.A.S., I.M.C., F.B.J., A.B.B., C.V. provided technical support, discussions and revisions; F.M.C. and J.J. co-wrote the manuscript and conceived the study; J.J. provided funding; J.J. provided supervision and revisions; All authors discussed the results and commented on and approved the manuscript.

## SUPPLEMENTAL INFORMATION

Extended Materials & Methods

Supplementary Figures 1-2

Supplementary Tables 1-2

